# Left Ventricular Geometry Improves Prediction of Sex-Specific Post-TAVR Remodeling in Aortic Stenosis

**DOI:** 10.64898/2026.03.30.26349680

**Authors:** Shoaib A. Goraya, Pauline Lauwers, Hoda Javadikasgari, Amir Rouhollahi, Ali Homaei, Shahab Masoumi, Edoardo Zancanaro, Mostafa Rezaeitaleshmahalleh, Brian C. Ayers, Sameer Hirji, Mohamad Alkhouli, Arminder Jassar, Iman Aganj, Ashraf Sabe, Farhad R. Nezami

**Affiliations:** Division of Cardiac Surgery, Brigham and Women’s Hospital, Boston, MA; Department of Surgery, Harvard Medical School, Boston, MA; Department of Information Technology and Electrical Engineering, ETH Zurich, Switzerland; Division of Cardiac Surgery, Massachusetts General Hospital, Boston, MA; Department of Cardiology, Mayo Clinic, Rochester, MN; Athinoula A. Martinos Center for Biomedical Imaging, Department of Radiology, Massachusetts General Hospital, Harvard Medical School, Charlestown, MA

**Keywords:** trans-aortic valve replacement, statistical shape analysis, left ventricular mass regression, machine learning, sex-specific shape modes

## Abstract

**Background:** Women with severe aortic stenosis (AS) are diagnosed later and experience poorer outcomes than men, partly because clinical approaches rely on 2D, valve-centric thresholds derived from male-predominant cohorts that underutilize information from 3D left ventricular (LV) geometry. We hypothesize that a sex-specific computational framework integrating statistical shape analysis (SSA) of pre-TAVR CT with machine learning would improve prediction of 1-year LV mass regression (LVMR).

**Objective:** To develop a computational framework leveraging 3D LV geometry and evaluate whether it improves sex-specific prediction of 1-year LVMR after TAVR.

**Methods:** We studied 339 patients with severe AS who underwent TAVR from 2013 to 2020 and had pre-TAVR CT and 1-year post-TAVR echocardiography. LV geometries were segmented into digital twins, and shape modes predictive of LVMR were extracted using SSA and partial least squares. These modes were incorporated into support vector regression models and compared with conventional echocardiographic predictors, including pre-TAVR LVEF, LVMI, and E/A ratio. Performance was assessed using RMSE and R^2^.

**Results:** After one year, 65% of patients showed positive LVMR, with median regression of approximately 10%; regression was significant overall and within each sex (*p* < 0.001) and similar between sexes (*p* = 0.99). Predictive shape modes differed by sex (*p* < 0.01), with women showing more localized variation and men broader geometric gradients. Sex-specific shape modes outperformed general modes and clinical metrics, particularly in women (R^2^ = 0.80, RMSE = 0.09 vs. R^2^ = 0.59, RMSE = 0.13; clinical-only baseline R^2^ = 0.16, RMSE = 0.22). In men, sex-specific modes also performed strongly (R^2^ = 0.89, RMSE = 0.08).

**Conclusion:** In severe AS, 3D LV geometry predicts post-TAVR reverse remodeling more accurately than conventional metrics and may improve risk stratification, particularly in women.

## Introduction

Aortic stenosis (AS) is the most prevalent valvular heart disease in developed countries and a major contributor to cardiovascular morbidity and mortality.^1–4^ Progressive calcification and stiffening of the aortic valve impose chronic pressure overload on the left ventricle (LV), triggering hypertrophy, fibrosis, and maladaptive remodeling that drive adverse outcomes.^5–8^ With an aging population, the burden of AS is expected to increase substantially.^2,9^ Transcatheter aortic valve replacement (TAVR) has become an established treatment for severe AS across the surgical risk spectrum.^10–13^Although valve replacement relieves afterload and promotes reverse LV remodeling in many patients, a substantial subset exhibits limited or adverse left ventricular mass regression (LVMR), which is associated with persistent dysfunction and worse clinical outcomes.^14–17^ Accurate prediction of postoperative remodeling is therefore critical for optimizing patient selection, procedural timing, and longitudinal management.

Current predictors of LV remodeling remain limited. Conventional metrics such as LV mass index, diastolic dysfunction, and ejection fraction do not leverage the complex three-dimensional structure of the LV available in contemporary imaging modalities such as computed tomography (CT).^18–20^ This limitation is particularly important because AS is not sex neutral.^21–27^ Women more often exhibit concentric remodeling, smaller LV cavities, higher filling pressures, and low-flow physiology, while men more commonly demonstrate eccentric remodeling and cavity dilation.^23–25,28^ These sex-related differences are inadequately captured by conventional, largely valve-centric clinical metrics in diagnostic pathways that are predominantly developed in male-dominant cohorts, creating an important gap in risk stratification and prediction of recovery after intervention (Figure 1).^22,23,29–35^

**Figure 1.**
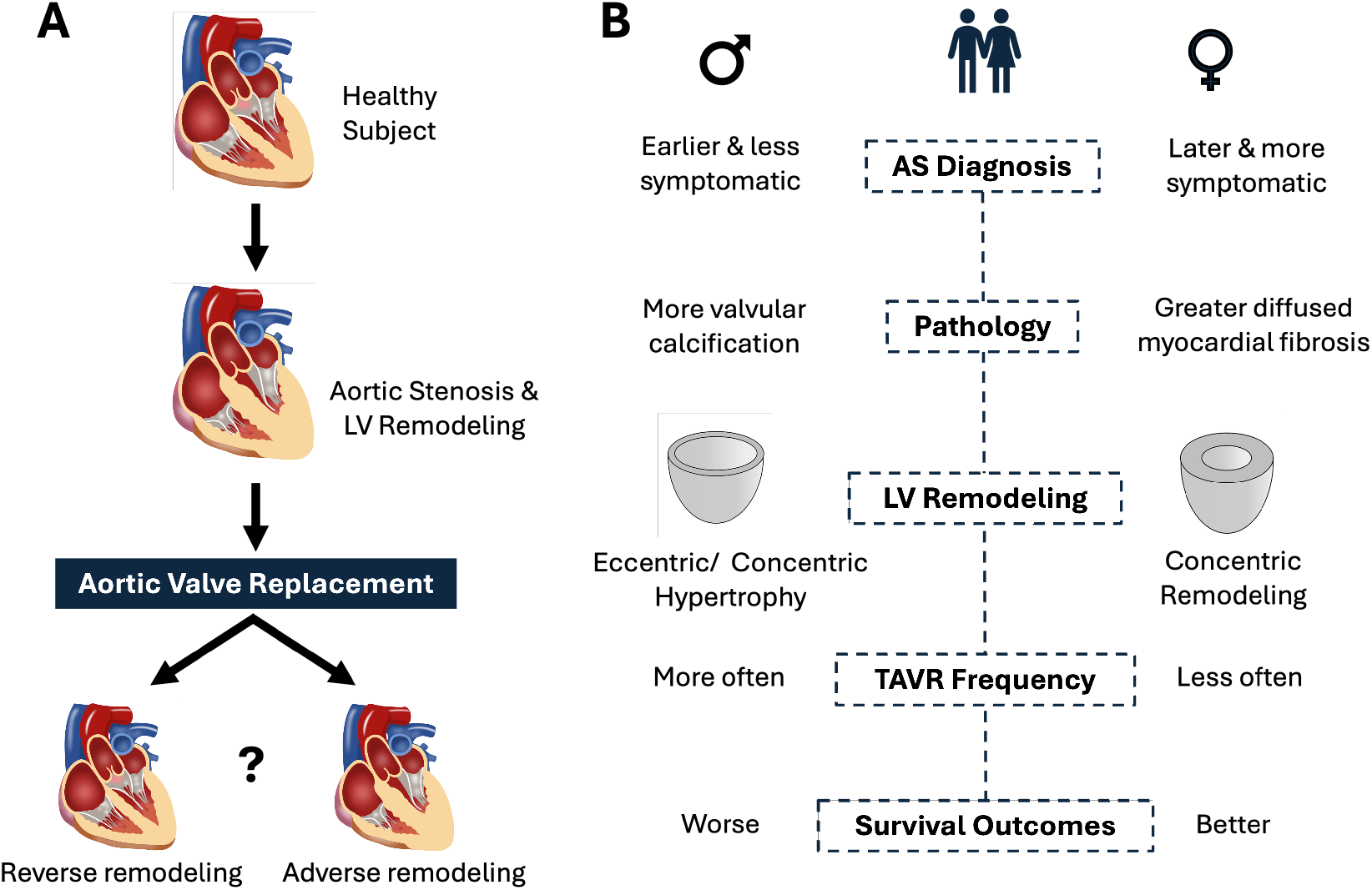
Aortic stenosis and sex-specific disparities. **(A)** Aortic stenosis is characterized by progressive calcium deposition on the valve leaflets, leading to narrowing of the aortic orifice, increased LV workload, and compensatory hypertrophy. Valve replacement can reverse or exacerbate LV remodeling, underscoring the need for accurate prediction of post-intervention outcomes to guide individualized treatment planning. **(B)** Sex-specific differences exist in AS pathophysiology, LV remodeling, and response to TAVR; although women often experience greater survival benefit following TAVR, they face delayed diagnosis, less frequent intervention, and have distinct patterns of LV remodeling compared with men.

Accordingly, we sought to identify sex-specific imaging markers of post-TAVR remodeling. We hypothesized that preoperative CT contains latent geometric information that can be quantified using statistical shape analysis (SSA), and that these geometric descriptors would improve prediction of postoperative remodeling in a sex-specific manner beyond conventional clinical and echocardiographic measures. By integrating computational modeling, dimensionality reduction, and advanced image analysis, SSA enables extraction of quantitative descriptors of LV geometry that may reveal sex-dependent structural determinants of reverse remodeling not accessible through standard metrics alone.

In this study, we developed a multidisciplinary computational pipeline (Figure 2) integrating AI-enabled CT reconstruction and SSA to characterize LV geometry and predict postoperative remodeling. Using a multicenter cohort of 339 patients with severe AS undergoing TAVR, we evaluated whether preoperative LV shape features improve prediction of reverse remodeling and clinical outcomes, with specific emphasis on sex-specific remodeling signatures. This framework establishes LV geometry as a sex-aware mechanistic imaging biomarker of cardiac recovery beyond conventional measures and provides a foundation for improved risk stratification, personalized therapeutic decision-making, and future optimization of transcatheter valve therapies in severe AS.

**Figure 2.**
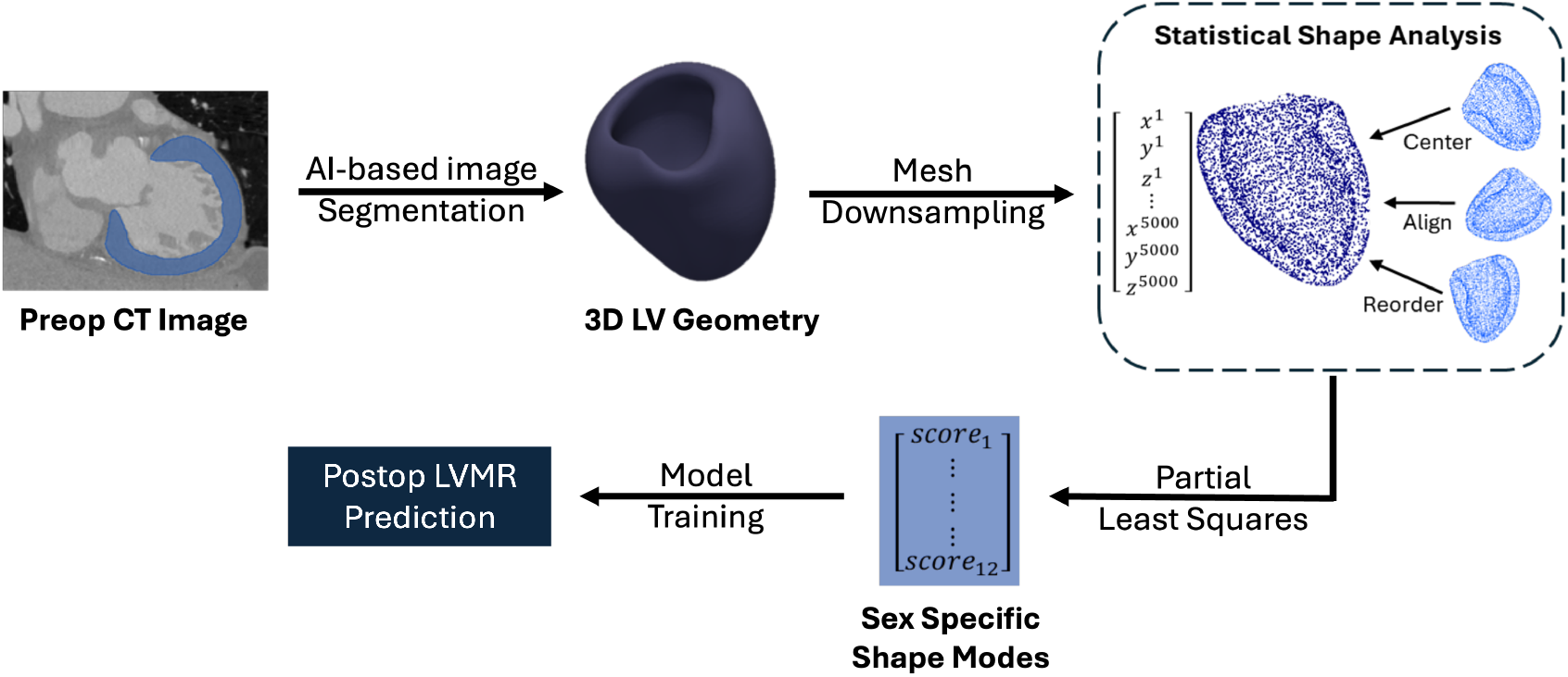
Leveraging 3D LV geometry and shape features to improve sex-specific prediction of post-TAVR LVMR. The pipeline begins with AI-based segmentation of pre-TAVR CT images to generate digital twin of 3D LV geometries. Statistical shape analysis and partial least square regression are performed on the digital twins to extract sex-specific shape modes that are most predictive of LV remodeling post-TAVR. These shape modes are then used to train support vector regression models to predict post-TAVR LVMR.

## Methods & Materials

### Study population

We retrospectively identified patients with severe aortic stenosis who underwent transcatheter aortic valve replacement (TAVR) at Brigham and Women’s Hospital and Massachusetts General Hospital (Boston, MA) between 2015 and 2023. Inclusion criteria were: (1) availability of a contrast-enhanced CT scan performed within 12 months prior to TAVR (±3 months); (2) a transthoracic echocardiogram (TTE) obtained within 7 days before or after the procedure; and (3) a follow-up TTE conducted approximately 12 months post-TAVR (±3 months). Patients who did not have a follow-up TTE were excluded. As a result, a total of 339 patients were included, of which 133 (39.2%) were women and 206 (60.8%) were men. Women were underrepresented among TAVR patients in our cohort (MGH: 34%; BWH: 41%), consistent with prior reports of gender-related differences in AS presentation and referral patterns^36^. Women often exhibit distinct valve and ventricular remodeling phenotypes that may delay recognition and timing of intervention, contributing to their lower representation even when applying neutral clinical inclusion criteria (severe AS, analyzable CT, longitudinal TTE)^37^.

### CT image segmentation and LVMI Extraction

Utilizing contrast-enhanced CT scans obtained at end-diastole, the TotalSegmentator, an open-source deep learning model validated for multi-organ segmentation on CT, was employed to segment the LV. Manual post-processing was conducted as necessary to ensure the segmented geometries were watertight. Subsequently, the segmented LV was transformed into an STL 3D surface mesh via the marching cubes algorithm, yielding approximately 300,000 geometric points per patient. The left ventricular mass index (LVMI) was determined from transthoracic echocardiograms (TTE) using the formula recommended by the American Society of Echocardiography (ASE)^38^ (see Supplemental Material).

To verify the accuracy of our CT-based LV segmentations, we computed several geometric measures, including LV mass, sphericity, wall thickness, endocardial cavity volume, and mass-to-volume ratio, for both women and men (Supplemental Table 1). These values were consistent with those reported in the literature.

LV mass regression was defined as:

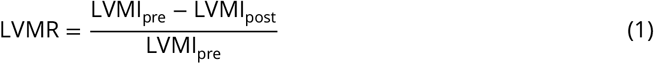

Preoperative and 1-year postoperative LVMI values were also extracted from echocardiography reports. LVMI measured from CT and echocardiography showed some differences (Supplemental Figure 2). These differences arise from (i) beam/plane positioning and foreshortening when probe orientation changes between scans, (ii) caliper/contour placement variability on 2D/M-mode/Doppler images, and (iii) intrinsic physiologic variability (arrhythmia, ventilatory effects)^39^. Echo is also an operator-dependent exam in which artifacts or omissions can occur^40^. Hand-held or focused cardiac ultrasound is primarily qualitative rather than quantitative, further limiting mass estimation^41^. Since echo is clinically ubiquitous, we used echo-based LV mass for longitudinal outcomes (pre- and post-TAVR LVMR) while extracting shape features from pre-TAVR CT; this modality mix is pragmatic but imperfect.

### Statistical shape analysis of 3D LV geometries

Each 3D left ventricular shape was first centered by translating its centroid to the coordinate origin. A reference anatomy was then selected as the shape whose surface area and volume were closest to the cohort’s average. All individual shapes were rigidly aligned to this reference using the Iterative Closest Point (ICP) algorithm^42^, which applies only rotations and translations to ensure an anatomically consistent orientation. For each subject, a deviation vector was obtained by subtracting the mean shape from the individual shape representation.

### Partial least square regression to extract LV shape modes

Shape features, derived from statistical shape analysis of 3D LV geometries, were used as predictors in a partial least squares (PLS) regression^43^ model to identify geometric patterns associated with LVMR. Instead of treating each point on the 3D surface separately, PLS summarizes thousands of anatomical measurements into a smaller set of “components,” each representing a pattern of geometric variation. These components can be thought of as the dominant shape features that best predict remodeling. As the number of PLS components increased, the cumulative variance in LVMR explained by the model (R^2^) increased steadily (Supplemental Figure 1). In the overall cohort, approximately 98% of the variance in LVMR was captured using 15 components. The female-only and male-only analyses required fewer components (11 and 13, respectively) to reach a similar level of explanatory power. In practical terms, this means that the retained components summarize nearly all predictive information contained in the 3D geometry while avoiding overfitting or unnecessary complexity (Supplemental Figure 1).

### Characterizing gender-specific dimorphism in LV remodeling using shape modes

To evaluate whether patterns of LV remodeling differ between men and women, we repeated the full PLS-based shape analysis separately in three groups: women only, men only, and the overall combined cohort. This allowed us to identify gender-specific shape modes and compare them with those derived from the entire study population.

Next, we examined whether the shape modes extracted from the full cohort contained any gender-specific information. For each mode, we compared the distribution of patient-specific scores between men and women using Welch’s two-sample t-test. To quantify the similarity or difference between the male- and female-derived modes, we computed a cross-similarity matrix based on the cosine similarity between the mode-loading vectors (see Supplemental Material). In cross-similarity matrix, values near 0 indicate nearly identical patterns of remodeling and values near 1 indicate markedly different geometric patterns.

To help visualize and interpret what each PLS mode represents anatomically, we used a raycasting technique (Supplemental Figure 3) that maps the LV shape into intuitive distance-based “heatmaps”. These distances formed two-dimensional maps showing the cavity size (endocardial distances) and wall thickness (difference between epicardial and endocardial distances). To interpret a specific mode, we generated shapes corresponding to a one-standard-deviation increase along that mode and compared their distance maps with those of the mean LV. The resulting difference maps highlight the regions where the cavity expands or contracts or where the wall thickness increases or decreases, thereby providing clinically interpretable visualizations of each remodeling pattern.

### Model training for prediction of post-TAVR LVMR

To develop a predictive model of post-TAVR LVMR based on pre-TAVR 3D LV geometries, we trained Support Vector Regression (SVR) on patient-specific shape scores obtained from the PLS analysis. SVR was selected because it can capture complex, non-linear relationships between LV remodeling and clinical outcomes. Model training and testing were performed using a shuffle–split cross-validation strategy repeated five times, where 20% of patients were randomly held out as an independent test set in each split.

Predictive performance was assessed using two standard regression matrices (see Supplemental Material). The coefficient of determination (R^2^) measures how well the model explains variation in observed LVMR, providing a direct measure of predictive accuracy. The root mean squared error (RMSE) was used to quantify the average magnitude of prediction errors. Because RMSE is expressed in the same units as LV mass index, it provides a clinically intuitive measure of how close the model’s predictions are to the true degree of remodeling.

## Results

### Baseline cohort characteristics

Baseline demographic, comorbidity, and echocardiographic characteristics are detailed in Table **??**. Clinical comorbidities, including diabetes, hypertension, and dialysis status, were extracted from electronic medical records. Baseline variables encompassed standard demographic factors such as age and body surface area, key medical conditions, and comprehensive echocardiographic assessments of cardiac structure and function. Echocardiographic parameters included septal and posterior wall thickness, left ventricular internal diameter at end-diastole, left ventricular mass and mass index, as well as indices of diastolic function, such as the E/A ratio and the presence or absence of diastolic dysfunction.

### Left ventricular mass regression post-TAVR

Preoperative and 1-year postoperative LVMI values were obtained from clinically reported echocardiography. Although LVMI derived from CT and echocardiography demonstrated some differences (Supplemental Figure 2), echocardiography-derived LVMI was used for longitudinal outcome assessment and prediction model given its routine clinical use and widespread availability. LV mass regression (LVMR) was calculated according to Eq. 1, representing the relative change in LVMI and quantifying the degree of reverse ventricular remodeling following TAVR. As shown in Figure 3, the majority of patients exhibited positive LVMR, reflecting a reduction in LV mass and favorable structural remodeling at 1 year post-TAVR. Overall, 65% of patients demonstrated positive LVMR (ΔLVMI>0), whereas 35% exhibited no regression or an increase in LVMI (ΔLVMI≤0); these proportions were similar between men and women. The distribution of LVMR, along with pre- and post-TAVR LVMI values, is shown in Figure 3. Median LVMI decreased significantly at 1 year compared with baseline in the overall cohort and within each sex (paired Wilcoxon signed-rank test, two-sided *p* < 0.001). The median relative regression was approximately 10% in the entire cohort, with comparable regression observed between men and women. No significant difference in LVMR was observed between men and women (*p* = 0.99).

**Figure 3.**
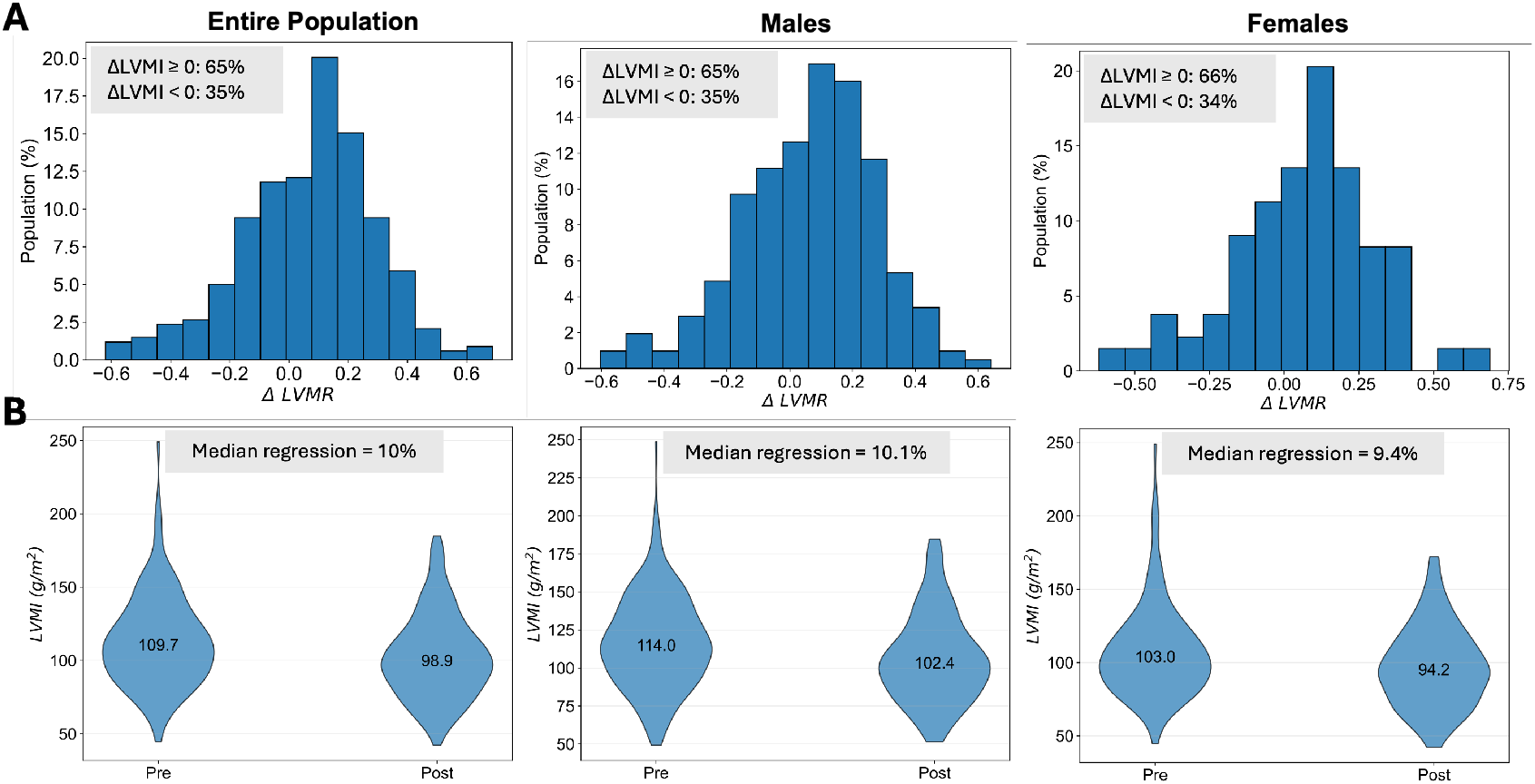
LV mass regression and pre-post changes assessed by echocardiography. **(A)** Histogram of changes in LVMI in the entire cohort (n=339), males (n=206), and females (n=133). 65% of patients demonstrated positive LVMI regression (ΔLVMI > 0), whereas 35% showed either no regression or an increase (ΔLVMI ≤ 0). These proportions were comparable when stratified by sex. **(B)** Violin plots of pre- and post-TAVR LVMI in the males, females, and the entire cohort, showing a significant regression in LV mass post-TAVR within each group (paired Wilcoxon signed-rank test, *p* < 0.001). Median regression between males and females were around 10% and not statistically significant (*p* = 0.99).

### Sex-specific analysis of shape modes extracted from 3D LV geometries

Shape modes predictive of post-TAVR LVMR were derived from 3D LV geometries using statistical shape analysis and partial least squares regression. Several modes differed significantly between sexes (Supplemental Figure 4), including Modes 1, 2, 4, 6, and 12 (*p* < 0.01). Women demonstrated higher scores for Modes 1, 4, and 6, whereas men demonstrated higher scores for Modes 2 and Ray-casting maps associated Modes 1, 2, and 12 with global geometric variation, including LV size and wall thickness, whereas Modes 4 and 6 reflected more localized changes in ventricular sphericity (Supplemental Figure 5).

Variable importance projection (VIP) analysis identified distinct anatomical regions associated with LVMR (Supplemental Figure 6). In women, influential regions were distributed across endocardial and epicardial surfaces, including the LV base. In men, influential regions were more localized, primarily at the basal LV. Sex-stratified mean geometries demonstrated a more spherical LV morphology in women and a more elongated geometry with thinner walls in men.

Cross-dissimilarity analysis demonstrated both shared and sex-specific remodeling modes (Figure 4). Using a predefined dissimilarity threshold (d ≥ 0.85), Female Modes 1–3 and Male Modes 5, 7, and 11 were identified as sex-specific. Female modes demonstrated more localized wall displacement, whereas male modes exhibited broader, more diffuse geometric variation (Figure 5 and Table 2). These findings demonstrate distinct sex-specific geometric phenotypes associated with post-TAVR remodeling.

**Figure 4.**
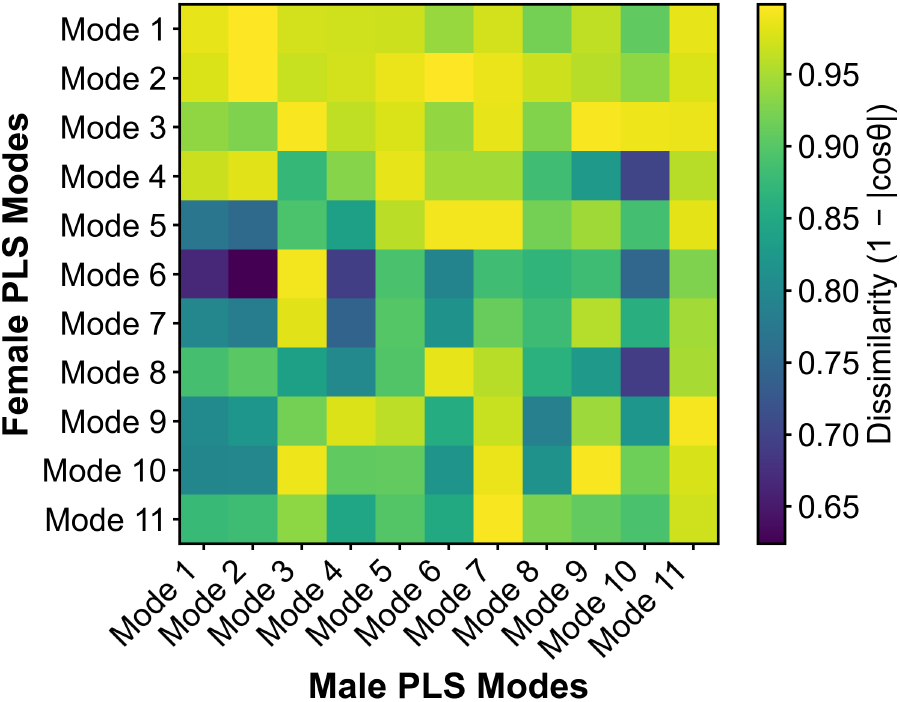
Cross-dissimilarity heatmap comparing female and male shape modes. Each cell shows the dissimilarity score between a female mode (rows) and a male mode (columns), where darker colors indicate higher similarity. Values near 0 indicate nearly identical patterns of remodeling and values near 1 indicate markedly different geometric patterns. Using a dissimilarity threshold of 85%, Female Modes 1, 2, and 3 and Male Modes 5, 7, and 11 emerged as sex-specific, indicating that dominant female modes are unique to women, while the primary male modes align with female counterparts and male-specific remodeling features appear only in higher-order modes.

**Figure 5.**
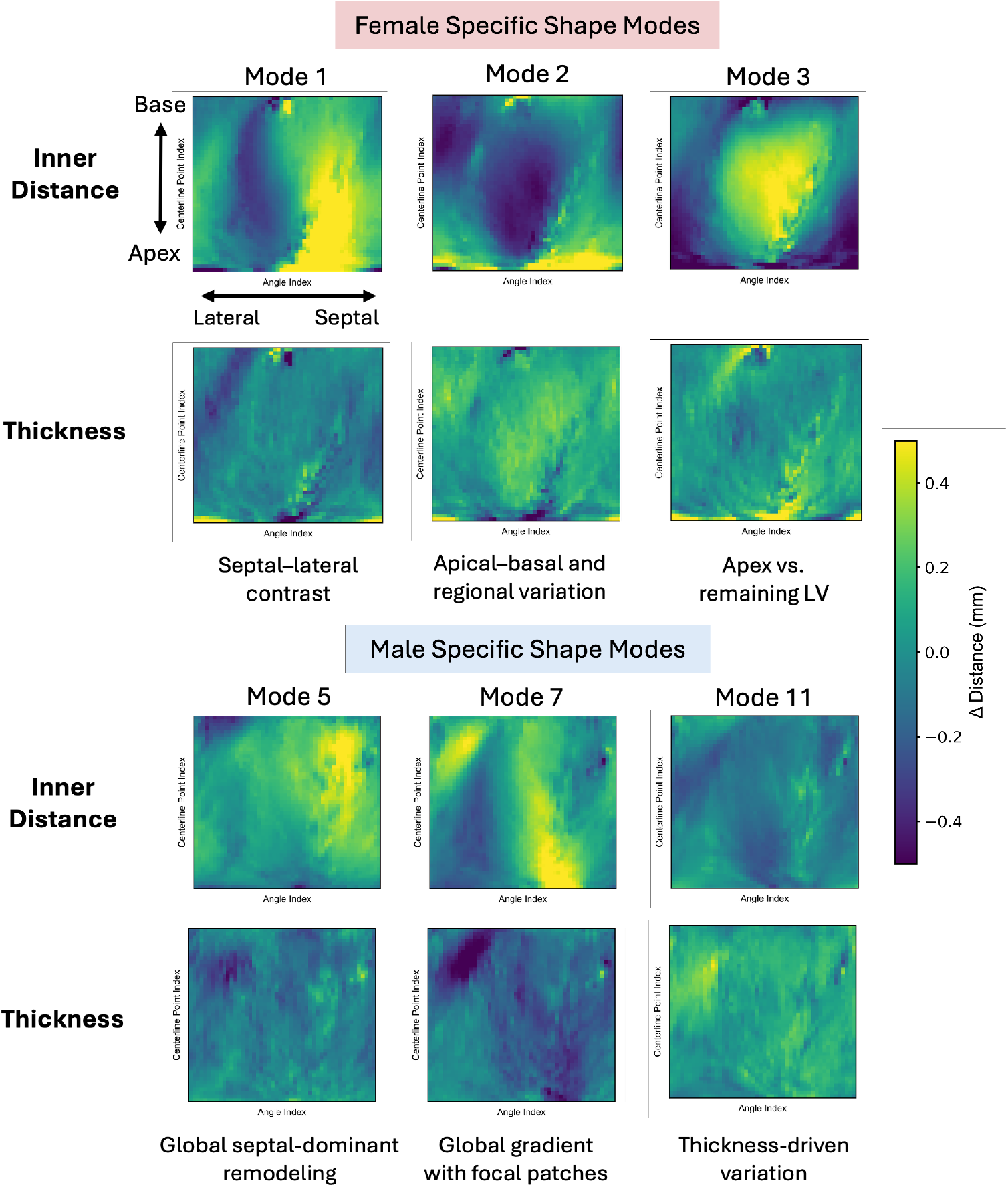
Heatmaps showing distinct LV remodeling in men and women. ΔDistance shows movement of LV wall in either inward (negative sign) or outward (positive sign) direction. Female modes (top) reveal localized remodeling patterns, such as apical and septal hotspots, whereas male modes (bottom) are characterized by broader global gradients with embedded regional patches. These complementary patterns highlight distinct geometric remodeling signatures between men and women.

**Table 1.** Baseline demographic, comorbidity, and echocardiographic variables stratified by sex.

| **Variable** | **N** | **Mean ± SD or N(%)** |
| --- | --- | --- |
| **Age** | **339** | **78.35 ± 8.52** |
| Male | 206 | 77.35 ± 8.41 |
| Female | 133 | 79.24 ± 8.54 |
| **Body Surface Area (m<sup>2</sup>)** | **253** | **1.94 ± 0.24** |
| Male | 150 | 2.02 ± 0.21 |
| Female | 103 | 1.82 ± 0.24 |
| **Diabetes** | **242** | **72 (29.8)** |
| Male | 144 | 44 (30.6) |
| Female | 98 | 28 (28.6) |
| **Hypertension** | **244** | **220 (90.2)** |
| Male | 145 | 130 (89.7) |
| Female | 99 | 90 (90.9) |
| **Current Dialysis** | **244** | **5 (2.0)** |
| Male | 145 | 2 (1.4) |
| Female | 99 | 3 (3.0) |
| **Interventricular Septum Thickness (cm)** | **226** | **1.05 ± 0.17** |
| Male | 135 | 1.06 ± 0.19 |
| Female | 91 | 1.03 ± 0.13 |
| **Left Ventricular Posterior Wall Thickness (cm)** | **199** | **1.01 ± 0.09** |
| Male | 124 | 1.02 ± 0.11 |
| Female | 75 | 1.00 ± 0.06 |
| **Left Ventricle Internal Diameter End Diastole (cm)** | **252** | **4.06 ± 0.81** |
| Male | 150 | 4.20 ± 0.78 |
| Female | 102 | 3.86 ± 0.82 |
| **Left Ventricular Mass (g)** | **253** | **198.44 ± 62.47** |
| Male | 150 | 213.12 ± 61.64 |
| Female | 103 | 177.07 ± 57.56 |
| **Left Ventricular Mass Index (g/m<sup>2</sup>)** | **339** | **113.11 ± 31.18** |
| Male | 206 | 116.82 ± 30.44 |
| Female | 133 | 107.36 ± 31.54 |
| **Left Ventricular Ejection Fraction (%)** | **226** | **59.58 ± 9.75** |
| Male | 134 | 58.59 ± 9.70 |
| Female | 92 | 61.03 ± 9.69 |
| **E/A Ratio** | **173** | **1.10 ± 0.74** |
| Male | 102 | 1.15 ± 0.86 |
| Female | 71 | 1.03 ± 0.54 |

**Table 2.** Anatomical interpretation of sex-specific shape modes describing distinct LV remodeling. Effects correspond to baseline +1 SD.

| Mode & sex | General pattern | Effect of +1 SD (inner-distance / thickness / spatial notes) |
| --- | --- | --- |
| Mode 1 (Female) | Septal-lateral contrast | Inner-distance decreases in septal/apical regions while relatively preserved in lateral/inferior walls; thickness remains largely unchanged. |
| Mode 2 (Female) | Apical-basal and regional variation | Inner-distance increases in lateral and anterior apical regions, while septal and inferior walls show decreases along most of the LV; mild overall thinning. |
| Mode 3 (Female) | Apex vs. remaining LV | Inner-distance increases at the apical tip, whereas most of the remaining LV walls exhibit decreased inner-distance; accompanied by mild thinning. |
| Mode 5 (Male) | Global septal-dominant remodeling | Smooth decrease in inner-distance across most walls, most pronounced in basal septal regions; thickness unchanged. |
| Mode 7 (Male) | Global gradient with focal patches | Broad gradient in inner-distance with focal effects in apical septum and basal lateral wall; thickness generally increases. |
| Mode 11 (Male) | Thickness-driven variation | Minimal net change in inner-distance; diffuse reduction in wall thickness. |

### Predictive modeling of post-TAVR LVMR

We evaluated a support vector regression (SVR) model to predict LVMR after TAVR using sex-specific shape scores as the predictors. In women, the SVR model built from female-specific shape modes demonstrated substantially better performance than models using general (population-derived) modes, achieving a cross-validated mean of R^2^ = 0.80 and RMSE = 0.09, compared with R^2^ = 0.59 and RMSE = 0.13, respectively (Figure 6). Predictions based on sex-specific modes closely matched the observed LVMR values, with less scatter and fewer systematic errors. Among men, sex-specific modes also yielded strong performance, with R^2^ = 0.89 and RMSE = 0.08 under the same evaluation framework.

**Figure 6.**
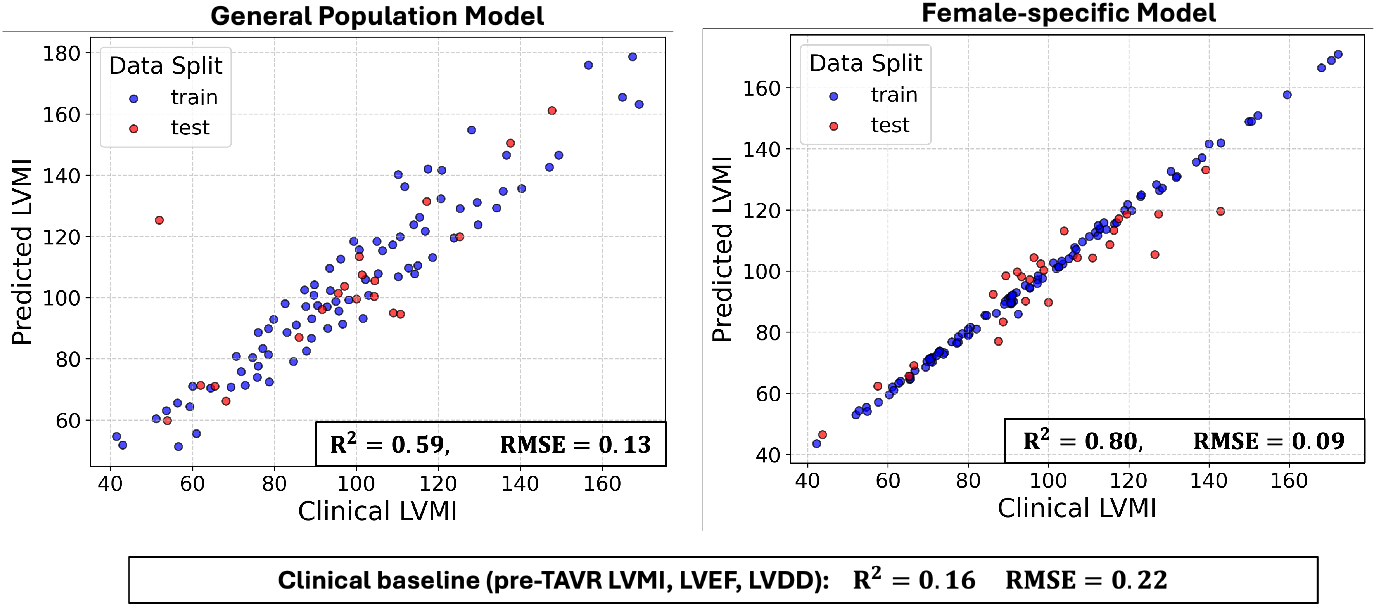
Predictive models of post-TAVR LVMR that are based on 3D LV shape modes significantly outperform clinical baseline model. **(A)** Predictive model using sex-specific shape modes outperform general population model. Models based on female-specific and male-specific shape modes achieved high predictive accuracy in women (R^2^ = 0.80) and men (R^2^ = 0.89), respectively. **(B)** Predictive performance using general population modes (non-sex-specific) decreased (R^2^ = 0.59) with more scatter and systematic deviations around clinical LVMI. As a comparison, the baseline clinical model using LVEF, LVDD, and LVMI as predictors of post-TAVR LVMR showed poor predictive ability (R^2^ = 0.16).

For comparison, we constructed a baseline clinical model using linear regression on three routinely available pre-TAVR echocardiographic variables: LV mass index (LVMI), left ventricular ejection fraction (LVEF), and diastolic dysfunction (LVDD). LVDD was classified based on a simplified E/A threshold (abnormal if *E*/*A* < 0.8 or *E*/*A* ≥ 2.0). This clinical-only model showed poor predictive ability, with *R*^2^ = 0.16 and RMSE = 0.22, underscoring the added value of detailed geometric shape descriptors in forecasting post-TAVR remodeling.

## Discussion

### Novel computational framework linking pre-TAVR LV geometry to post-TAVR remodeling in severe aortic stenosis

The key finding of this study is that pre-TAVR LV geometry on routine CT carries clinically meaningful information about the likelihood of reverse remodeling after TAVR. Using a computational framework based on 3-dimensional shape analysis, we identified geometric patterns associated with subsequent LV mass regression that were not captured by conventional measures alone. These findings suggest that the ventricle retains a structural imprint of chronic pressure overload and myocardial adaptation before intervention, and that this information may help explain why some patients recover more favorably than others after valve replacement.

From a clinical perspective, these results support CT-based LV geometry as a potential imaging biomarker for improved risk stratification in severe AS. Because CT is already routinely obtained before TAVR, this approach could be incorporated into existing workflows without requiring additional imaging. Identifying patients who are less likely to undergo favorable reverse remodeling may improve patient selection, inform postprocedural monitoring, and support personalized management strategies beyond conventional echocardiographic indices which do not fully capture the structural determinants of ventricular recovery.

### Shape modes from 3D LV geometries are sex-differentiating

A second key finding is that reverse remodeling after TAVR appears to follow distinct geometric patterns in women and men. In women, the LV features linked to subsequent LV mass regression were more localized, with focal regional changes carrying much of the predictive signal. In men, the relevant geometric changes were more diffuse and distributed across the ventricle. Thus, the remodeling signal in women was concentrated in a smaller number of dominant features, whereas in men it was spread across a broader set of geometric characteristics. These findings suggest that women and men may not follow the same pathway of myocardial adaptation and recovery in severe aortic stenosis. Clinically, this is important because it implies that a single, sex-neutral imaging model may miss important determinants of recovery after TAVR. A sex-aware approach may therefore improve prediction of reverse remodeling, help refine postprocedural expectations, and support more individualized follow-up and management. Importantly, these results highlight the value of 3D shape analysis for revealing biologically and clinically relevant differences that are not apparent from conventional imaging metrics.

### Sex-specific prediction models of post-TAVR LVMR outperform conventional clinical metrics

A third key finding is that sex-specific models based on pre-TAVR LV geometry substantially out-performed conventional clinical measures for predicting LV mass regression after TAVR. Standard echocardiographic parameters, including baseline LV mass index, ejection fraction, and LV dimension, showed limited predictive value (*R*^2^ = 0.16), whereas CT-derived 3D LV geometry markedly improved prediction (*R*^2^ = 0.59). Performance improved further with sex-stratified models, both in women (*R*^2^ = 0.80) and in men (*R*^2^ = 0.89), highlighting that the relationship between ventricular structure and recovery after TAVR is inherently sex specific.

These findings have direct clinical relevance. Current risk assessment relies largely on global and valve-centric measures that offer limited insight into myocardial recovery after intervention. In contrast, sex-specific geometric phenotyping may help identify patients who are more or less likely to undergo favorable reverse remodeling and guide more individualized follow-up after the procedure. More broadly, these results support the incorporation of sex-aware, CT-based structural markers into prognostic assessment beyond conventional imaging metrics alone.

### Clinical perspective: Can sex-specific LVMR prediction improve TAVR patient outcomes?

Timing of aortic valve intervention is one of the greatest challenges in AS, particularly in asymptomatic patients. Findings from our study extend beyond just the prediction of LV mass regression. Identification of adverse geometric phenotypes prior to TAVR may improve risk stratification and enable recognition of patients at risk of persistent ventricular dysfunction despite relief from valvular obstruction. Such information could enhance patient counseling, inform post-procedural monitoring strategies, and potentially support consideration of earlier intervention before irreversible remodeling develops. Because preprocedural CT imaging is routinely obtained for TAVR planning, this approach leverages existing clinical data without additional imaging burden. With automated segmentation and standardized shape modeling, integration of geometry-based biomarkers into existing clinical workflows appears technically feasible, with potential for reproducible implementation across centers. If prospectively validated, this framework may support scalable implementation across structural heart programs and facilitate incorporation of sex-informed risk assessment into clinical decision-making.

## Study Limitations

The outcome measure in our predictive model was echocardiographic LVMI. Although echocardiography is widely used in clinical practice, it remains operator dependent and derives LV mass from 2-dimensional measurements, making it vulnerable to variability in image acquisition and measurement.^39,40,44^In contrast, CT and CMR provide volumetric LV mass assessment with greater reproducibility and closer agreement with each other.^45,46^In our cohort, Deming regression showed both constant and proportional bias between echocardiographic and CT-derived LV mass, supporting CT as a more robust anatomic reference. Because pre-TAVR CT and echocardiography were not always performed on the same day, interval remodeling due to severe AS may also have contributed to cross-modality differences. Ideally, LV mass would be measured longitudinally with the same volumetric modality, but routine post-TAVR CT is not standard practice. We therefore used echocardiographic LVMI for follow-up remodeling assessment and CT-derived shape features for baseline phenotyping, and our findings should be interpreted in light of these modality-specific differences.

## Conclusion

In severe aortic stenosis, pre-TAVR 3D left ventricular geometry provides prognostic information on post-intervention reverse remodeling. Sex-specific shape analysis identified distinct remodeling phenotypes and improved prediction of LV mass regression beyond conventional metrics, supporting a role for sex-aware computational imaging in risk stratification and timing of intervention.

## Data Availability

All data produced in the present study are available upon reasonable request to the authors.

## Acknowledgment

Computing resources were provided by the NSF-ACCESS program MDE250010. This support is gratefully acknowledged.

## References

1. Iung B, Delgado V, Rosenhek R, et al. Contemporary presentation and management of valvular heart disease: the EURObservational Research Programme Valvular Heart Disease II Survey. Circulation. 2019;140:1156–1169.

2. Nkomo VT, Gardin JM, Skelton TN, Gottdiener JS, Scott CG, and Enriquez-Sarano M. Burden of valvular heart diseases: a population-based study. Lancet. 2006;368:1005–1011.

3. Michalski B, Dweck MR, Marsan NA, et al. The evaluation of aortic stenosis, how the new guidelines are implemented across Europe: a survey by EACVI. Eur Heart J Cardiovasc Imaging. 2020;21:357–362.

4. Ramos J, Monteagudo JM, Gonzalez-Alujas T, et al. Large-scale assessment of aortic stenosis: facing the next cardiac epidemic? Eur Heart J Cardiovasc Imaging. 2018;19:1142–1148.

5. Sacks MS and Yoganathan AP. Heart valve function: a biomechanical perspective. Philos Trans R Soc Lond B Biol Sci. 2007;362:1369–1391.

6. Levy D, Garrison RJ, Savage DD, Kannel WB, and Castelli WP. Prognostic implications of echocardiographically determined left ventricular mass in the Framingham Heart Study. N Engl J Med. 1990;322:1561–1566.

7. Une D, Mesana TG, Chan V, et al. Clinical impact of changes in left ventricular function after aortic valve replacement: analysis from 3112 patients. Circulation. 2015;132:741–747.

8. Fuchs C, Mascherbauer J, Rosenhek R, et al. Gender differences in clinical presentation and surgical outcome of aortic stenosis. Heart. 2010;96:539–545.

9. Lindroos M, Kupari M, Heikkilä J, and Tilvis R. Prevalence of aortic valve abnormalities in the elderly: an echocardiographic study of a random population sample. J Am Coll Cardiol. 1993;21:1220–1225.

10. Hirji S, Trager L, Harloff M, et al. Surgical Aortic Valve Replacement Outcomes in Non–Transcatheter Aortic Valve Replacement Centers: Implications for Tier-Based Systems of Care. Ann Thorac Surg. 2022;113:66–74.

11. Kawano Y, Newell P, Harloff M, et al. Early outcomes of transatrial mitral valve replacement in severe mitral annular calcification. JTCVS Techniques. 2021;9:49–56.

12. Harloff M, Papoy A, Hirji S, et al. Aortic root replacement to accommodate future valve-in-valve transcatheter aortic valve replacement. Ann Thorac Surg. 2021;111:e437–e438.

13. Percy E, Harloff M, Hirji S, et al. Nationally representative repeat transcatheter aortic valve replacement outcomes: report from the Centers for Medicare and Medicaid Services. JACC Cardiovasc Interv. 2021;14:1717–1726.

14. Heusch G, Libby P, Gersh B, et al. Cardiovascular remodelling in coronary artery disease and heart failure. Lancet. 2014;383:1933–1943.

15. Coronel R, Wilders R, Verkerk A, Wiegerinck R, Benoist D, and Bernus O. Electrophysiological changes in heart failure and their implications for arrhythmogenesis. Biochim Biophys Acta Mol Basis Dis. 2013;1832:2432–2441.

16. Levy D, Garrison R, Savage D, Kannel W, and Castelli W. Prognostic implications of echocar-diographically determined left ventricular mass in the Framingham Heart Study. N Engl J Med. 1990;322:1561–1566.

17. Azevedo P, Polegato B, Minicucci M, Paiva S, and Zornoff L. Cardiac remodeling: concepts, clinical impact, pathophysiological mechanisms and pharmacologic treatment. Arq Bras Cardiol. 2015;106:62–69.

18. O’Connor SA, Morice MC, Gilard M, et al. Revisiting sex equality with transcatheter aortic valve replacement outcomes: a collaborative, patient-level meta-analysis of 11,310 patients. J Am Coll Cardiol. 2015;66:221–228.

19. Treibel T, Badiani S, Lloyd G, and Moon J. Multimodality imaging markers of adverse myocardial remodeling in aortic stenosis. JACC Cardiovasc Imaging. 2019;12:1532–1548.

20. Gastl M, Behm P, Haberkorn S, et al. Role of T2 mapping in left ventricular reverse remodeling after TAVR. Int J Cardiol. 2018;266:262–268.

21. Pawade T, Clavel MA, Tribouilloy C, Dreyfus J, Mathieu P, Tastet L, et al. Computed Tomography Aortic Valve Calcium Scoring in Patients With Aortic Stenosis. Circ Cardiovasc Imaging. 2018;11:e007146.

22. Shan Y and Pellikka PA. Aortic stenosis in women. Heart. 2020;106:970–976.

23. Iribarren AC, AlBadri A, Wei J, et al. Sex differences in aortic stenosis: Identification of knowledge gaps for sex-specific personalized medicine. Am Heart J Plus. 2022;21:100197.

24. Tastet L, Kwiecinski J, Pibarot P, Capoulade R, Bürgi S, Nsaibia MJ, et al. Sex-Related Differences in the Extent of Myocardial Fibrosis in Patients With Aortic Stenosis. JACC Cardiovasc Imaging. 2020;13:1997–2008.

25. Singh A, Greenwood JP, Berry C, Shah ASV, McCann GP, Greenwood JP, et al. Sex differences in left ventricular remodelling, myocardial fibrosis and outcomes in aortic stenosis. Heart. 2019;105:1818–1826.

26. Tribouilloy C, Bohbot Y, Rusinaru D, Belkhir K, Diouf M, Altes A, et al. Excess Mortality and Undertreatment of Women With Severe Aortic Stenosis. J Am Heart Assoc. 2021;10:e018816.

27. Rice CT, Barnett S, O’Connell SP, et al. Impact of gender, ethnicity and social deprivation on access to surgical or transcatheter aortic valve replacement in aortic stenosis: a retrospective database study in England. Open Heart. 2023;10:e002373.

28. Appleby C, Bleiziffer S, Bramlage P, Delgado V, Eltchaninoff H, Gebhard C, et al. Sex-related disparities in aortic stenosis from disease awareness to treatment: a state-of-the-art review. J Thorac Dis. 2024;16:6308–6319.

29. Nguyen V, Mathieu T, Melissopoulou M, et al. Sex differences in the progression of aortic stenosis and prognostic implication: the COFRASA-GENERAC study. JACC Cardiovasc Imaging. 2016;9:499–501.

30. Cramariuc D, Rieck Å, Staal E, et al. Factors influencing left ventricular structure and stress-corrected systolic function in men and women with asymptomatic aortic valve stenosis (a SEAS Substudy). Am J Cardiol. 2008;101:510–515.

31. Simard L, Cote N, Dagenais F, et al. Sex-related discordance between aortic valve calcification and hemodynamic severity of aortic stenosis: is valvular fibrosis the explanation? Circ Res. 2017;120:681–691.

32. Tribouilloy C, Bohbot Y, Rusinaru D, et al. Excess mortality and undertreatment of women with severe aortic stenosis. J Am Heart Assoc. 2021;10:e018816.

33. Onorati F, D’Errigo P, Barbanti M, et al. Different impact of sex on baseline characteristics and major periprocedural outcomes of transcatheter and surgical aortic valve interventions: results of the multicenter Italian OBSERVANT Registry. J Thorac Cardiovasc Surg. 2014;147:1529– 1539.

34. Chaker Z, Badhwar V, Alqahtani F, et al. Sex differences in the utilization and outcomes of surgical aortic valve replacement for severe aortic stenosis. J Am Heart Assoc. 2017;6:e006370.

35. Iribarren A, AlBadri A, Wei J, et al. Sex differences in aortic stenosis: Identification of knowledge gaps for sex-specific personalized medicine. Am Heart J Plus. 2022:100197.

36. Vahanian A, Beyersdorf F, Praz F, et al. 2021 ESC/EACTS Guidelines for the management of valvular heart disease. Eur Heart J. 2022;43:561–632.

37. Shan Y and Pellikka PA. Aortic stenosis in women. Heart. 2020;106:970–976.

38. Treibel TA, Badiani S, Lloyd G, and Moon JC. Multimodality Imaging Markers of Adverse Myocardial Remodeling in Aortic Stenosis. JACC Cardiovasc Imaging. 2019;12:1532–1548.

39. Jozwiak M, Mercado P, Teboul JL, and Monnet X. What is the lowest change in cardiac output that transthoracic echocardiography can detect? Critical Care. 2019;23:116.

40. Wiegers SE, Ryan T, Arrighi JA, et al. 2019 ACC/AHA/ASE Advanced Training Statement on Echocardiography (Revision of the 2003 ACC/AHA Clinical Competence Statement on Echocardiography): A Report of the ACC Competency Management Committee. J Am Coll Cardiol. 2019;74:377– 402.

41. Spencer KT, Kimura BJ, Korcarz CE, Pellikka PA, Rahko PS, and Siegel RJ. Focused Cardiac Ultrasound: Recommendations from the American Society of Echocardiography. J Am Soc Echocardiogr. 2013;26:567–581.

42. Besl PJ and McKay ND. “Method for registration of 3-D shapes”. Sensor Fusion IV: Control Paradigms and Data Structures. Vol. 1611. Spie. 1992, pp. 586–606.

43. Wold S, Sjöström M, and Eriksson L. PLS-regression: a basic tool of chemometrics. Chemom Intell Lab Syst. 2001;58:109–130.

44. Lang RM, Badano LP, Mor-Avi V, et al. Recommendations for Cardiac Chamber Quantification by Echocardiography in Adults: An Update from the American Society of Echocardiography and the European Association of Cardiovascular Imaging. J Am Soc Echocardiogr. 2015;28:1– 39.

45. Grebe SJ, Malzahn U, Donhauser J, et al. Quantification of left ventricular mass by echocardiography compared to cardiac magnetic resonance imaging in hemodialysis patients. Cardiovasc Ultrasound. 2020;18:39.

46. Han D, Shanbhag A, Miller RJH, et al. AI-Derived Left Ventricular Mass From Noncontrast Cardiac CT: Correlation With Contrast CT Angiography and CMR. JACC Adv. 2024;3:101249.

