## Supplemental for "Left Ventricular Geometry Improves Prediction of Sex-Specific Post-TAVR Remodeling in Aortic Stenosis"

### Supplemental Material

#### Partial least squares to extract 3D LV shape features associated with post-TAVR LVMR

Shape features, derived from statistical shape analysis on 3D LV geometries, were used as predictors in a partial least squares (PLS) regression model to identify geometric patterns associated with LVMR. As the number of PLS components increased, the cumulative variance in LVMR explained by the model ($R^2$) increased steadily (Figure 1). In the overall cohort, approximately 98% of the variance in LVMR was captured using 15 components. The female-only and male-only analyses required fewer components (11 and 13, respectively) to reach a similar level of explanatory power. Across the groups, a small subset of components contributed most strongly to the model performance. In women, Components 1–3 had the greatest influence on the LVMR prediction. In men, components 3–5 had the greatest impact. In the full cohort, components 3, 5, and 6 had the largest marginal contributions. These findings suggest that although the overall remodeling patterns are similar across genders, the dominant geometric features associated with LVMR differ between men and women.

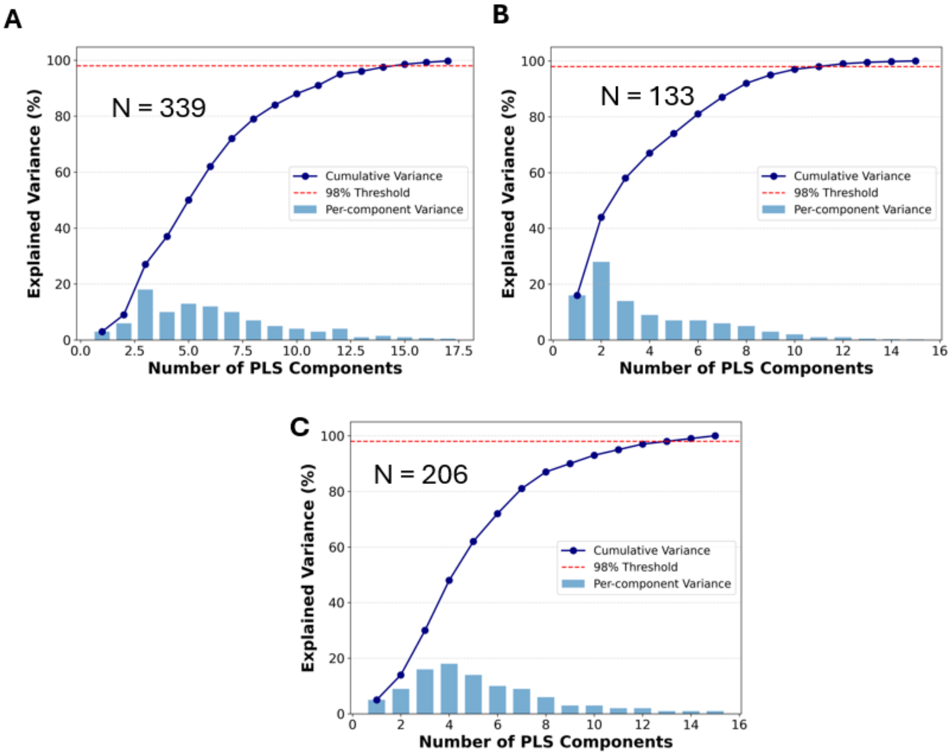


**Supplemental Figure 1. Explained variance of LVMR as a function of the number shape components derived through PLS of 3D LV geometries.** Cumulative variance explained in the outcome (LVMR) increases with the number of PLS components. To reach the 98% threshold, 16 components were required in the entire population, 11 in women, and 13 in men.

#### CT vs echo derived LV mass

The left ventricular mass index (LVMI) was determined from transthoracic echocardiograms (TTE) using the formula recommended by the American Society of Echocardiography (ASE)[38]:

$$\text{LV mass (g)} = 0.8 \times \left\{1.04 \times \left[(\text{IVSd} + \text{LVIDd} + \text{PWTd})^3 - (\text{LVIDd})^3\right]\right\} + 0.6 \tag{2}$$

$$\text{LVMI (g/m}^2) = \frac{\text{LV mass (g)}}{\text{BSA (m}^2)} \tag{3}$$

where *IVSd* is interventricular septal thickness at end-diastole (cm), *LVIDd* is the LV internal diameter at end-diastole (cm), *PWTd* is posterior wall thickness at end-diastole (cm), and *BSA* is body surface area ($m^2$).

Comparison between CT-derived and echocardiographic LV mass is shown in Figure 2. The Deming regression fit yielded a slope of $1.6$ and intercept of $-70$. The identity crossover occurs at $\approx 117$ g, so Echo underestimates CT below this value and increasingly overestimates CT above it.

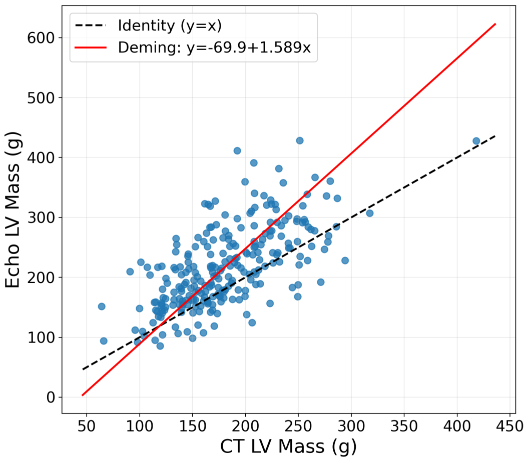


**Supplemental Figure 2. CT-derived vs echocardiographic LV mass.** Deming regression analysis on the LV mass computed from Echo and CT showed both proportional and constant bias. The black dashed line indicates identity ($y = x$) while the red line represents the Deming regression fit. Each dot represents one patient.

#### CT derived measurements

Segmentation-derived LV parameters differed between genders (Table 1). Men had greater absolute LV mass and endocardial volume, while women exhibited slightly higher sphericity and mass-to-volume ratio.

**Supplemental Table 1.** Gender-based comparison of segmented CT geometries.

| Variable | Women | Men |
|---|---|---|
| LV mass (g) | 146 | 194 |
| Sphericity | 0.45 | 0.43 |
| Thickness (mm) | 10.15 | 10.92 |
| Endocardial capped cavity volume (mL) | 72.26 | 96.32 |
| Mass(capped)/Volume (g/mL) | 1.33 | 1.29 |

#### Quantifying gender similarity in shape modes of men and women

To quantify the similarity of shape modes across genders, we computed the cosine of the angle between male and female mode pairs. Let $\mathbf{u}_i \in \mathbb{R}^p$ denote the female mode-$i$ loading vector and $\mathbf{v}_j \in \mathbb{R}^p$ the male mode-$j$ loading vector. The cosine similarity is defined as

$$\cos\theta_{ij} = \frac{\mathbf{u}_i^\top \mathbf{v}_j}{\|\mathbf{u}_i\|_2 \, \|\mathbf{v}_j\|_2},$$

and the corresponding dissimilarity index as

$$\text{Dissimilarity}_{ij} = 1 - \left|\cos\theta_{ij}\right|,$$

with values in $[0, 1]$, where $0$ indicates identical (or sign-flipped) directions and $1$ indicates orthogonality. Cosine dissimilarity was chosen over Pearson correlation because it compares the orientation of mode vectors in the full feature space, remaining scale- and sign-invariant while preserving baseline offsets. Unlike Pearson, which centers the data and can obscure such effects, cosine dissimilarity is more sensitive to directional misalignments that capture true geometric differences between modes. Consequently, it exposes sharper divergences between male and female loadings that Pearson may obscure. In practice, this yields a more reliable measure of whether two modes encode the same underlying deformation direction (irrespective of magnitude or sign), rather than merely indicating whether they impose similar subject rankings.

#### Ray casting approach for anatomical interpretation of LV shape modes

To interpret the anatomical deformation associated with individual PLS shape modes, we employed a ray-casting approach adapted to the elongated geometry of the left ventricle (Figure 3). Analogous to cylindrical coordinates, but accounting for the fact that the LV is not perfectly cylindrical, we first extracted a patient-specific centerline and used it as the effective z-axis for mapping. Along this axis, a fixed number of sampling points were defined, and at each location multiple radial rays were emitted in predefined angular directions within a plane perpendicular to the centerline. Each ray extended outward from the centerline until intersecting the endocardial or epicardial surface, and the traveled distance was recorded. Collectively, these measurements formed two-dimensional distance maps, where each pixel encodes the radial distance at a given axial and angular position. For mode interpretation, representative shapes were generated by perturbing the mean LV shape by one standard deviation along each PLS mode. The corresponding distance maps were then compared with those of the mean shape to obtain mode-specific difference heatmaps, highlighting regions of cavity expansion or contraction. The same procedure was applied to paired endocardial–epicardial intersections to construct myocardial thickness maps, thereby localizing regional differences in wall thickness associated with each PLS mode.

To quantify each mode systematically, we applied the ray-casting pipeline to two meshes per mode: (i) the population mean shape and (ii) the mean shape deformed by $+1$ SD along the mode. Point clouds were converted to watertight surfaces using the preprocessing. Analogous to cylindrical coordinates, but accounting for the fact that the LV is not perfectly cylindrical, we first extracted a patient-specific centerline and used it as the effective z-axis for mapping. Along this axis, a fixed number of sampling points were defined, and at each location multiple radial rays were emitted in predefined angular directions within a plane perpendicular to the centerline. Each ray extended outward from the centerline until intersecting the endocardial or epicardial surface, and the traveled distance was recorded. Collectively, these measurements formed two-dimensional distance maps, where each pixel encodes the radial distance at a given axial and angular position. Ray casting was run identically on both meshes to produce inner-distance and wall-thickness heatmaps.

Figure 3 illustrates the procedure. On the left is the baseline LV shape reconstructed from the population mean, and in the middle the same shape deformed by +1 SD along mode 1. Orange arrows indicate example ray paths used for distance mapping. The right panel shows the resulting heatmap of inner-distance differences (mode 1 vs. baseline), plotted as a function of centerline position (apex to base, y-axis) and angular orientation (lateral, inferior, septal, anterior, x-axis). Positive values (yellow) indicate inward displacement, while negative values (blue) indicate outward displacement.

The difference of the maps were then computed to isolate the modal effect.

$$\Delta d_{\text{in}} = d_{\text{in}}^{\text{deformed}} - d_{\text{in}}^{\text{baseline}}, \qquad \Delta w = w^{\text{deformed}} - w^{\text{baseline}}.$$

A positive $\Delta d_{\text{in}}$ means the deformed endocardium lies farther from the centerline (outward shift), while $\Delta w$ highlights regions of thickening ($\Delta w > 0$) or thinning ($\Delta w < 0$). This provides an anatomical, spatial description of each abstract mode rather than just a numeric vector. Because sampling is performed in a consistent centerline–azimuth coordinate system, heatmap features directly localize to septal, lateral, anterior, and inferior walls in apical or basal region. As a sanity check, repeating the procedure with a $-1$ SD deformation produced sign-reversed difference maps, confirming internal consistency and reproducibility.

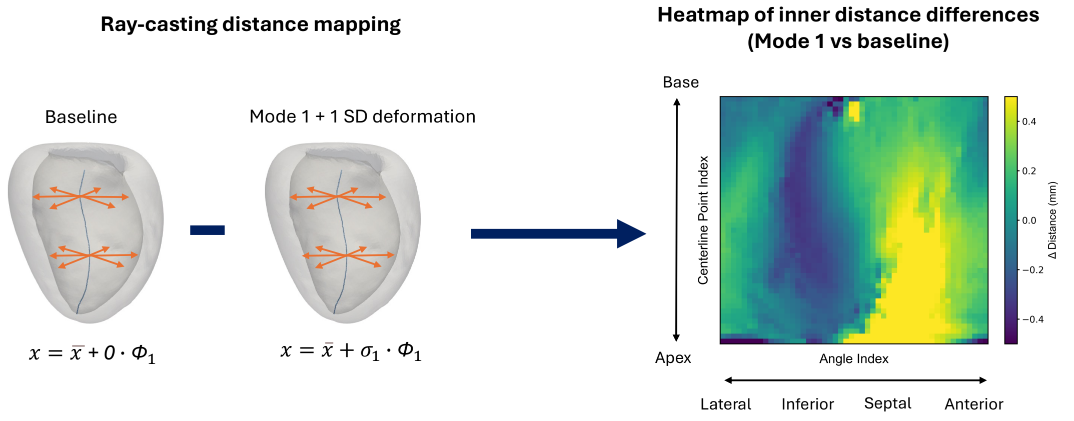


**Supplemental Figure 3. Ray-casting–based quantification of LV shape mode deformation.** On the left, baseline left ventricular shape reconstructed from the population mean. In the middle, shape deformed along female mode 1 by +1 standard deviation (SD). Orange arrows indicate example ray paths used for distance mapping. On the right heatmap of inner distance differences between baseline and deformed shapes, plotted as a function of centerline position (apex to base, y-axis) and angular orientation (lateral, inferior, septal, anterior, x-axis). Positive values (yellow) indicate inward displacement, while negative values (blue) indicate outward displacement. This approach enables systematic quantification of subtle geometric changes associated with each shape mode.

#### Gender-specifc differences in shape modes derived from 3D LV geometries

In the shape modes derived from 3D LV geometries of general population, several of them showed statistically significant differences in shape scores between women and men (Figure 4). Specifically, sape modes 1, 2, 4, 6, and 12 differed by gender with $p < 0.01$ (two-sided group comparison). Men exhibited higher scores for Modes 2 and 12, whereas women had higher scores for Modes 1, 4, and 6.

To physiologically interpret these gender-specific modes, we adopted the ray-casting procedure as described previously. The resulting heatmaps indicate how each ventricular region is modified by a given mode (Figure 5). Qualitatively, ray-casting maps associated Modes 1, 2, and 12 with more global remodeling including LV size and wall thickness, while Modes 4 and 6 displayed more localized patterns in the form of spherical to ovoidal ratio.

#### Variable importance projection (VIP) analysis

VIP analysis was used to localize shape landmarks that most strongly contribute to the output variable (LVMR) in the PLS shape modes. By convention, $\text{VIP} > 1$ denotes above–average influence; for visualization we set model–specific thresholds to highlight exactly three hotspots per gender-specific model and 6 hotspots for the general population: entire population $\tau = 1.85$, women $\tau = 2.22$, and men $\tau = 2.00$ (Figure 6). In women, VIP hotspots appeared on the outer wall, inner wall, and at the base; in men, all three clustered basally; in the general model, hotspots were distributed across two endocardial and four epicardial sites. These regional patterns align with the gender–stratified mean shapes, where women exhibit a more rounded (higher sphericity) LV, men a more elongated and thinner geometry, and the general population mean lies intermediate.

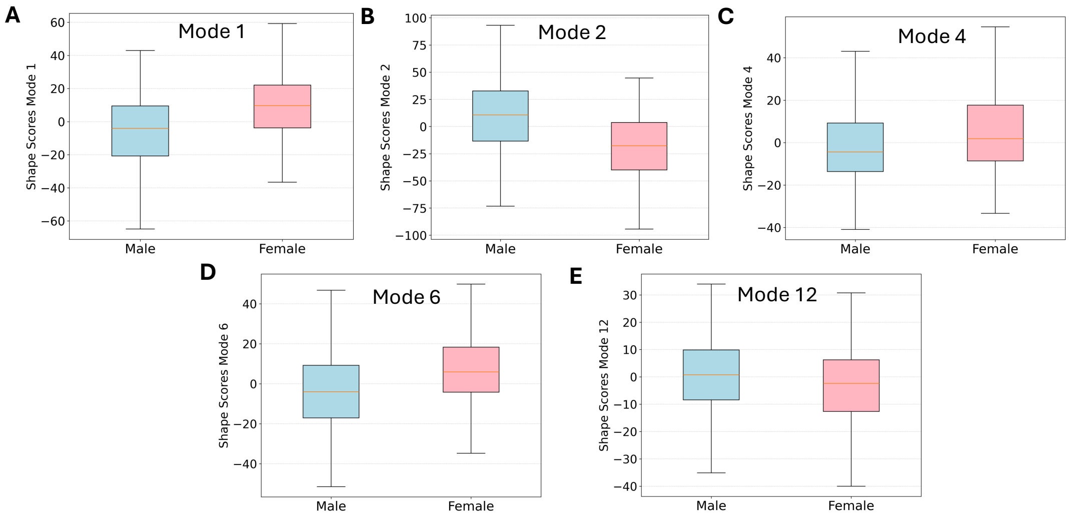


**Supplemental Figure 4. General shape modes differing by gender.** From the general modes on the entire population, not gender specific, Modes 1, 2, 4, 6 and 12 show statistically significant differences in shape scores between women and men with p-value < 0.01.

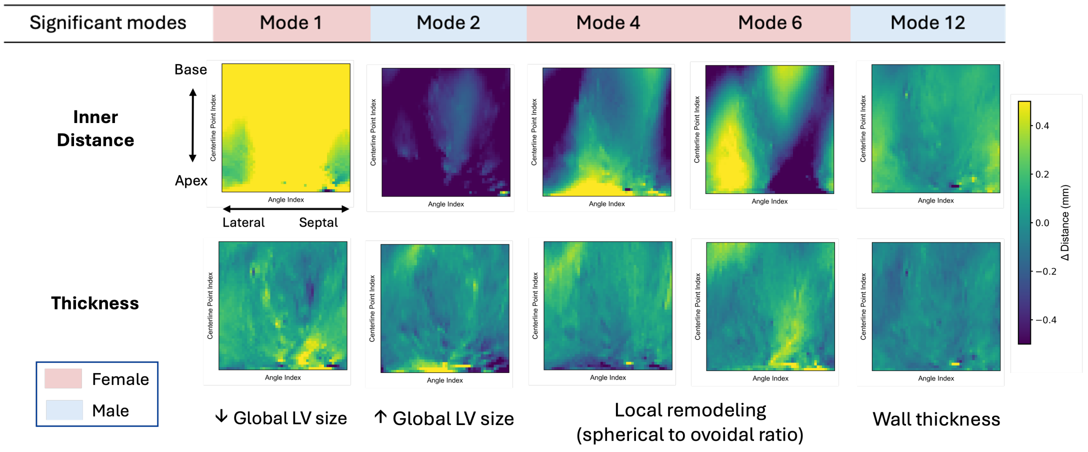


**Supplemental Figure 5. Heatmaps of general shape modes showing statistically significant differences in shape scores between men and women along with their anatomical explanation** (Welch's t-test, p < 0.05). Modes 1, 2, and 12 show more global remodeling gradients, while Modes 4 and 6 display more localized regional patterns.

#### Assessing predictive performance of models for LVMR prediction

Predictive performance was assessed using two standard regression metrics. The coefficient of determination ($R^2$) quantifies the proportion of variability in observed LV mass regression explained by the model:

$$R^2 = 1 - \frac{\sum_{i=1}^{n}(y_i - \hat{y}_i)^2}{\sum_{i=1}^{n}(y_i - \bar{y})^2},$$

where $y_i$ denotes the observed LVMI regression, $\hat{y}_i$ the model predictions, and $\bar{y}$ the sample mean. An $R^2$ of 1 indicates perfect prediction, 0 indicates that the model performs no better than predicting the mean value, and negative values indicate worse performance than that baseline.

The root mean squared error (RMSE) was used to quantify the average magnitude of prediction errors:

$$\text{RMSE} = \sqrt{\frac{1}{n}\sum_{i=1}^{n}(y_i - \hat{y}_i)^2}.$$

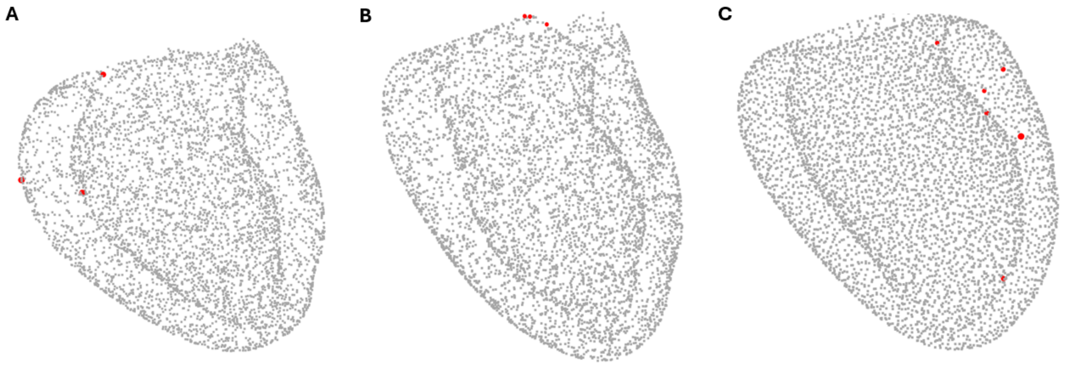


**Supplemental Figure 6. Visualization of anatomical regions that drive LV mass regression using variable importance projection (VIP) analysis. (A)** In women, the most influential regions (marked in red) were located on both the endocardial and epicardial surfaces and at the LV base. **(B)** In men, influential regions were concentrated primarily at the base. **(C)** In the combined cohort, VIP hotspots appeared at two endocardial and four epicardial regions.

---

Because RMSE is expressed in the same units as LV mass index, it provides a clinically intuitive measure of how close the model's predictions are to the true degree of remodeling.
